# Application of Machine Learning in Prediction of COVID-19 Diagnosis for Indonesian Healthcare Workers

**DOI:** 10.1101/2021.10.15.21265021

**Authors:** Shreyash Sonthalia, Muhammad Aji Muharoom, Sinta Amalia Kusumastuti Sumulyo, Fariza Zahra Kamilah, Fatma Aldila, Bijak Rabbani, Andhika Tirtawisata, Olivia Herlinda, Jatin Khaimani, Levana Sani, Astrid Irwanto, Rebriarina Hapsari, Nurul Luntungan, Diah Saminarsih, Akmal Taher

## Abstract

The COVID-19 pandemic poses a heightened risk to health workers, especially in low- and middle-income countries such as Indonesia. Due to the limitations of implementing mass RT-PCR testing for health workers, high-performing and cost-effective methodologies must be developed to help identify COVID-19 positive health workers and protect the spearhead of the battle against the pandemic. This study aimed to investigate the application of machine learning classifiers to predict the risk of COVID-19 positivity (by RT-PCR) using data obtained from a survey specific to health workers. Machine learning tools can enhance COVID-19 screening capacity in high-risk populations such as health workers in environments where cost is a barrier to the accessibility of adequate testing and screening supplies. We built two sets of COVID-19 Likelihood Meter (CLM) models: one trained on data from a broad population of health workers in Jakarta and Semarang (full model) and tested on the same, and one trained on health workers from Jakarta only (Jakarta model) and tested on both the same and an independent population of Semarang health workers. The area under the receiver-operating-characteristic curve (AUC), average precision (AP), and the Brier score (BS) were used to assess model performance. Shapely additive explanations (SHAP) were used to analyse future importance. The final dataset for the study included 5,393 healthcare workers. For the full model, the random forest was selected as the algorithm choice. It achieved cross-validation of mean AUC of 0.832 ± 0.015, AP of 0.513 ± 0.039, and BS of 0.124 ± 0.005, and was high performing during testing with AUC and AP of 0.849 and 0.51, respectively. The random forest classifier also displayed the best and most robust performance for the Jakarta model, with AUC of 0.856 ± 0.015, AP of 0.434 ± 0.039, and BS of 0.08 ± 0.0003. The performance when testing on the Semarang healthcare workers was AUC of 0.745 and AP of 0.694. Meanwhile, the performance for Jakarta 2022 test set was an AUC of 0.761 and AP of 0.535. Our models yielded high predictive performance and can be used as an alternative COVID-19 methodology for healthcare workers in Indonesia, therefore helping in predicting an increased trend of transmission during the transition into endemic.

## INTRODUCTION

Since the first confirmed case of COVID-19 in Indonesia in March 2020, there have been over 6.736 million confirmed cases and more than 160 thousand deaths resulting from COVID-19 infection as of March 1^st^, 2023 [1]. Healthcare workers in Indonesia are at a high risk of COVID-19 exposure and infection due to the nature of the profession, with 2,087 COVID-19-related deaths as of March 1^st^, 2023 [2,3]. Although the government had undergone several efforts to prevent any further expansion of the spread of the virus, such as strengthening testing, tracing, and treatment [4], as of March 2023, Indonesia is ranked as the eleventh country with the biggest total deaths caused by the virus according to WHO COVID-19 Dashboard [5]. Furthermore, due to the often-asymptomatic nature of the infection [6], efforts to prevent COVID-19 transmission were constrained by the ability to immediately detect and isolate the infected people [7–9].

The study was conceived in 2021 as an initial response to the surge in positive cases and resource constraints for mass reverse transcription transcription-polymerase chain reaction (RT-PCR) testing. Mass testing by RT-PCR, the gold standard of COVID-19 diagnostic testing, remains one of the key measures to reduce transmission of the virus [10]. However, the implementation of mass RT-PCR testing is limited in developing countries such as Indonesia due to financial, capital, and logistical constraints [11,12]. Despite the rapidly increasing COVID-19 cases since the Eid al-Fitr holiday in May 2021, some regions of Indonesia have been facing limited reagent supply and inadequate laboratory capacity to provide sufficient testing. In June 2021, Indonesia had the second-lowest testing rate in Southeast Asia with only 7.5 tests per confirmed case, far below the World Health Organization (WHO) recommendation of 10-30 tests per confirmed case [13–15]. The healthcare system of Indonesia has further been weakened by surges in case numbers and patients requiring hospitalization, leading to depleted medical supplies [16]. Furthermore, in the event of a surge, one study assessed that several provinces in Indonesia would likely have suboptimal diagnostic capabilities even if using rapid diagnostic technologies in referral hospitals [17].

The most used screening method in Indonesia is temperature check using a digital thermometer, which only had a sensitivity of 9.9% according to a study conducted in Uganda [18]. The limitations of mass RT-PCR implementation in Indonesia underscore the development of a COVID-19 detection method that is accurate and can be accessible to users with minimum equipment or resources. Machine learning tools can achieve these goals and have already shown promise in several countries such as the United States, China, and Slovenia [19–22]. Some models have already been generated for COVID-19 screening using data from sources such as Computed Tomography (CT) scans [19,23], clinical symptoms [20–22], and laboratory tests [23–26]. The sensitivity and specificity values of these tools are high, ranging from 0.86-0.93 and 0.56-0.98 respectively. In Indonesia, the development of a COVID-19 screening tool using machine learning has been carried out, coined Corona Likelihood Metric 1.0 (CLM 1.0). CLM 1.0 was high-performing, with an accuracy of 78% (precision = 0.79, F1 score = 0.74), and was used as a method to detect COVID-19 for the mass population in DKI Jakarta. CLM 1.0 was a survey consisting of questions about symptomatology, demographics, and behavioral tendencies.

Currently, most machine-learning COVID-19 screening tools have been deployed in technologically advanced countries [19,20,23,25,27]. Since most of the available models used data from hospitalized patients, the tools may not be effective for COVID-19 screening for health workers, due to differences in available features between hospitalized patients and health workers. Hospitalized patients are highly likely to have different symptomatology, behavioral tendencies, and PPE usage requirements than health workers who are not hospitalized, the target population of this study. We aim to apply machine learning algorithms to develop software tools to augment COVID-19 screening for Indonesian health workers. This method is expected to ease the burden in Indonesia’s healthcare system caused by COVID-19 through the implementation of a fast, accessible, and widespread screening methodology that can allow for accurate triage and systematic allocation of RT-PCR testing for health workers.

## METHODS

### Ethics approval and consent to participate

Institutional Review Board (IRB) approval was granted by the Institute of Research and Community Service of Universitas Katolik Indonesia Atma Jaya (Jakarta, Indonesia) under the IRB Reference Number of 62A/III/LPPM.PM.10.05/05/2020. This study did not include minors. Written informed consents were obtained from respondents before they were enrolled in the study.

### Study design and data

The study was designed as a cross-sectional observational study to test the performance of five machine learning models in predicting COVID-19 positivity with a total of 5,393 health workers, including medical and non-medical professionals working at healthcare facilities, to test the performance of five machine learning models. The study was reported as per the requirement of the STROBE guidelines (S1 File). The data was collected between 20 January 2021 and 30 June 2022 in 103 healthcare facilities: 21 healthcare providers in the Greater Jakarta Area and West Java, and one testing facility in Semarang which covers 80 healthcare providers. The hospitals and healthcare providers were selected through online recruitment, recommendations from medical associations, and a partnership agreement with the following inclusion criteria: 1) the ability to conduct swab collection for RT-PCR testing by trained health workers, 2) the presence of healthcare or non-healthcare staff with COVID-19 symptoms or close contact with COVID-19 patients, and 3) the support from the health facility management to participate in the research. The proportion of respondents from Jakarta and West Java was 4,820 (89.4%), while from Semarang was 573 (10.6%).

Initially, the hospitals had the authority to prioritize their staff to be included in the study by following these criteria: either (1) had close contact with at least one COVID-19 patient within the last fourteen days or developed COVID-19-related symptoms within the last fourteen days and (3) consented to participate in the study. Meanwhile, individuals below 18 years old were excluded. However, during the surge of asymptomatic cases caused by the Delta and Omicron variants, the inclusion criteria were expanded to include all healthcare workers who were interested in participating in the study in exchange for a free COVID-19 RT-PCR test. For each respondent, we collected oropharyngeal or nasopharyngeal swab specimens for RT-PCR as well as data for symptoms, comorbidities, COVID-19 protective behaviors, working condition, and COVID-19 vaccination status through a self-administered questionnaire written in Indonesian. However, the authors were unable to access the personal information of each participant due to the data that has been deidentified by the testing labs. All swab specimens from respondents in the Greater Jakarta and West Java were tested at Primajaya Hospital, East Bekasi, while specimens from respondents in participating healthcare providers in Semarang were tested at Diponegoro National Hospital.

### Study Variables

The survey questions included demographics, behavioral (protective, social, and travel) tendencies, COVID-19 vaccination status, working conditions, symptoms, comorbidities, and level of COVID-19 exposure and interaction with infected patients at health facilities (S1 File). The dependent variable in this study was the result of the COVID-19 RT-PCR. Respondents with inconclusive RT-PCR results were not included in the processed dataset. Behavioral questions were chosen based on general and medical worker-specific risk factors identified in the current literature and encompassed handwashing, mask-wearing, PPE adherence, social distancing, and domestic and foreign travel tendencies. Hand-washing behaviors were assessed by the level of adherence to the six-step handwashing protocol [26,28]. Mask-wearing and social distancing behaviors were assessed according to current WHO guidelines [29, 30].

### Modelling and Prediction

To predict COVID-19 diagnosis in our cohort, we trained and evaluated several machine learning classification algorithms, including logistic regression, random forest [31], extra trees classifier [32], and model ensembling. These were implemented using the scikit-learn Python library [33], while XGBoost was implemented using scikit-learn compatible packages in Python. These models were chosen after experiments with various algorithms, such as optimization and deep learning methods, during preliminary modelling. Pre-processed respondent features were used as inputs for each model, generating an output prediction risk score with a value between 0 and 1. The output was then converted to a class label by a thresholding function. Hyperparameters were tuned and chosen using the random search optimization method in scikit learn [35]. Feature selection was implemented using the sequential feature selection method in scikit learn. Model performance was analyzed and interpreted using the area under the receiving-operating characteristic curve (AUC) and area under the precision-recall (PR) curve, average precision (AP), while model calibration was assessed using the Brier Score (BS) for each model. Feature importance and model interpretability was assessed using Shapley additive explanations (SHAP) from the SHAP package [36] in Python.

Two sets of models were developed: one using respondents from health facilities in both Jakarta and Semarang (full model) and one on respondents from Jakarta health facilities only (Jakarta model). The full model was trained with stratified 5-fold cross-validation on 80% of the dataset and tested on the remaining 20%. The Jakarta model was trained only using data from Jakarta in 2021 and was tested on inputs from the Semarang health workers as well as the data from Jakarta in 2022. This was developed to assess the predictive capability of this approach in an independent population and to evaluate the model persistency in different periods and see if there is a significant difference in its performance. Due to a class imbalance between positive and negative classes in the dataset as shown in Table 1, the model was trained using sample weights to give more exposure to the minority (positive) class. This was done through the hyperparameters *class_weight* for random forest, extra trees classifier, logistic regression, and *scale_pos_weight* for XGBoost.

**Table 1.**
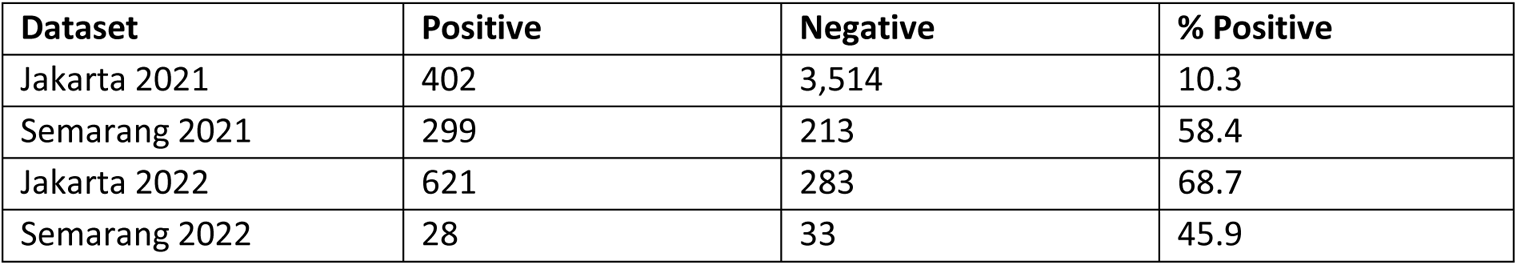
Summary of test results.

| Dataset | Positive | Negative | % Positive |
| --- | --- | --- | --- |
| Jakarta 2021 | 402 | 3,514 | 10.3 |
| Semarang 2021 | 299 | 213 | 58.4 |
| Jakarta 2022 | 621 | 283 | 68.7 |
| Semarang 2022 | 28 | 33 | 45.9 |

## RESULTS

### Data Summary

Data from 5,393 healthcare workers were included in the final dataset for model building. A summary of cohort demographic is provided in Table 2. Approximately 74.5% of respondents were females and the average age was 30.5 years old. There was no missing data for each variable of interest as all questions in the questionnaire were set to be mandatory As health workers are the priority group for receiving the COVID-19 vaccine in Indonesia, 89.17% of respondents had received at least the first dose of the COVID-19 vaccine. Regarding protective behaviors in the prior month, most respondents indicated they had been trained in PPE standards (98.13%) and six-step hand washing techniques (97.70%). Around 18.99% of respondents were currently self-isolating after having close contact with COVID-19 patients and 30.82% were involved in aerosol-generating procedures on COVID-19 patients. Furthermore, approximately half of positive cases were currently self-isolating. Additionally, around 75.43% and 62.3% of respondents reported having activity in a closed room and having outdoor activities at least 1-3 times a month, respectively. This study also found that 74.87% of respondents always wore a mask outside the home, 51.07% always avoided shaking hands, and 50.34% always maintained physical distance. In terms of the use of public transportation, 79.73% of respondents never used mass public transportation and 73.37% never used door to door transportation.

**Table 2.**
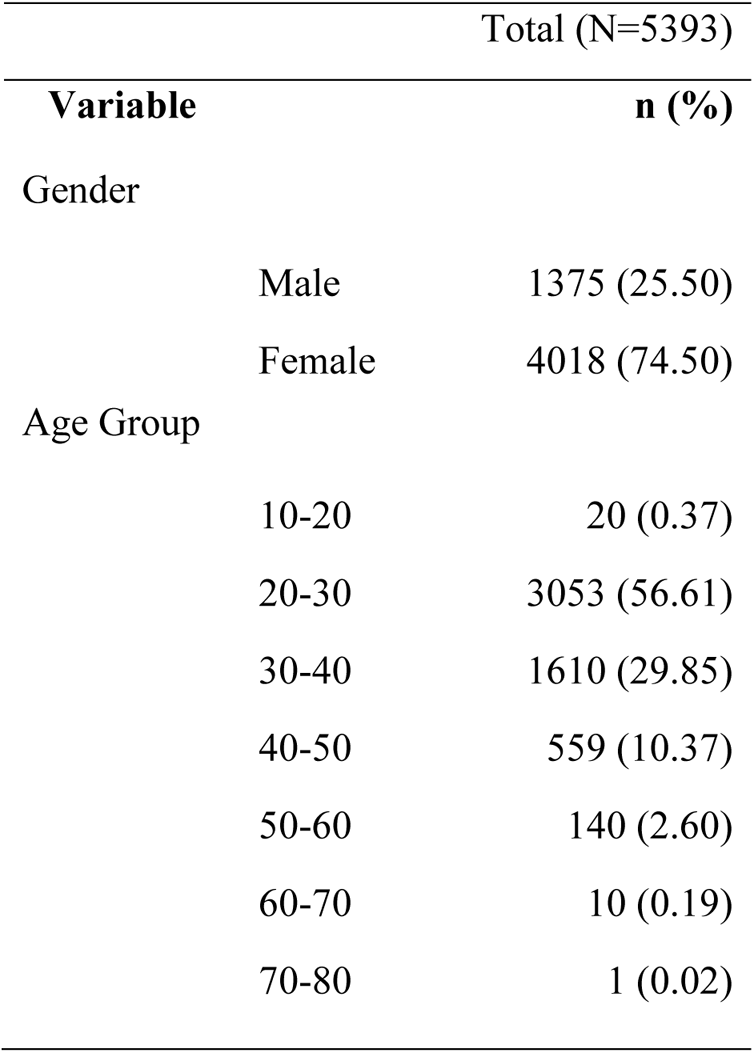
Demographic data of 5,393 healthcare workers

Approximately 80% of positive cases were symptomatic. Cough (53.23%), headache (46.11%), runny nose (42.76%), sore throat (38.13%), and myalgia (36.95%) were the five most reported symptoms among those who had COVID-19 positive test results. The proportion of 90.82% of respondents stated that they did not have any comorbidities. Pregnancy (3.13%), lung disease (2.38%), and hypertension (2.16%) were the most reported comorbidities among those who were COVID-19 positive.

### Model Performance and Explainability

The full model contains data from both Jakarta and Semarang, and thus is a model on a wider population and would be generalizable to a more heterogenous testing population. Meanwhile, the Jakarta model was developed to investigate the model performance on a geographically independent test set, which was the data from the Semarang health workers, and a different time period, which was the data from Jakarta in 2022.

#### Full Model

Regarding the performance of the model, Figure 1 displays the validation set ROC and PR curves as well as training set calibration curves for all the algorithms applied to the full patient cohort training set. The prevalence of positive classes in the validation and testing datasets was 17.2%. The best performing model during 5-fold stratified cross-validation was the extra tree classifier and random forest classifier with an average AUC (0.832 ± 0.015), followed by XGBoost classifier (0.830 ± 0.018), voting classifier (0.826 ± 0.017), and the logistic regression (0.812 ± 0.015) (Fig. 1A). The XGBoost classifier also had the best AP (0.513 ± 0.047) followed by that of the random forest (0.514 ± 0.04), voting classifier (0.511 ± 0.0334), and logistic regression (0.492 ± 0.04) (Fig. 1B). The training set calibration curves showed random forest, voting classifier, and extra trees were well-calibrated, while the logistic regression and XGBoost overpredicted risk for COVID-19 infection in the bottom half of bins and had the highest brier scores (0.168 ± 0.007 and 0.204 ± 0.005) (Fig. 1C). Random forest had the lowest Brier score (0.124 ± 0.005), followed by voting classifier (0.142 ± 0.007) and extra trees classifier (0.154 ± 323 0.005).

**Fig. 1.**
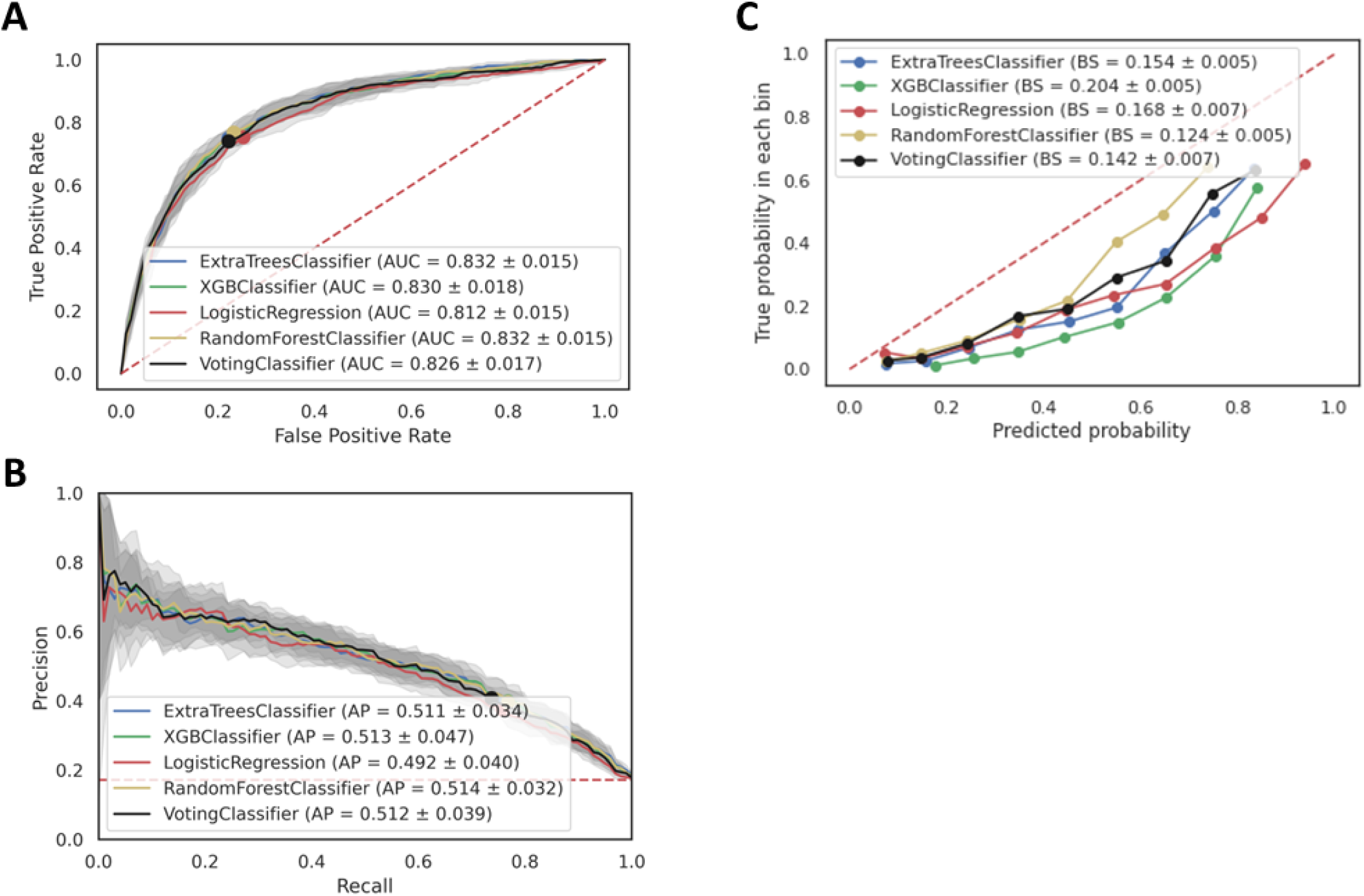
Full model cross validation results. **A,** ROC curve on the validation sets for all classifiers. **B,** PR curve on validation sets for all classifiers. **C,** Calibration curve for all classifiers generated from validation sets during 5-fold cross validation.

On the held-out test set, random forest produced the highest average AUC of 0.849 as compared to voting classifier (0.848), extra tree classifier (0.847), the XGBoost (0.846), and logistic regression (0.839) (Fig. 2A). The random forest classifier had the highest AP (0.51) while extra tree, XGBoost, voting classifier, and logistic regression had AP of 0.509, 0.503, 0.497, and 0.487 respectively (Fig. 2B). Using operating thresholds from the cross-validation step, we could calculate the recall, specificity, positive-predictive value (PPV), negative-predictive value (NPV), and the F1 score of our models (Fig. 2C). In this perspective, the the voting classifier had the highest F1 score (0.565) and was followed by extra tree classifier (0.562) in second place. Throughout the evaluation process, random forest produced the most stable results by either taking the first or the second place among the models that we applied. Furthermore, random forest also had the lowest Brier Score, which shows that the model has the best probability predictions. Hence, we picked random forest as our main model for the full model.

**Fig. 2.**
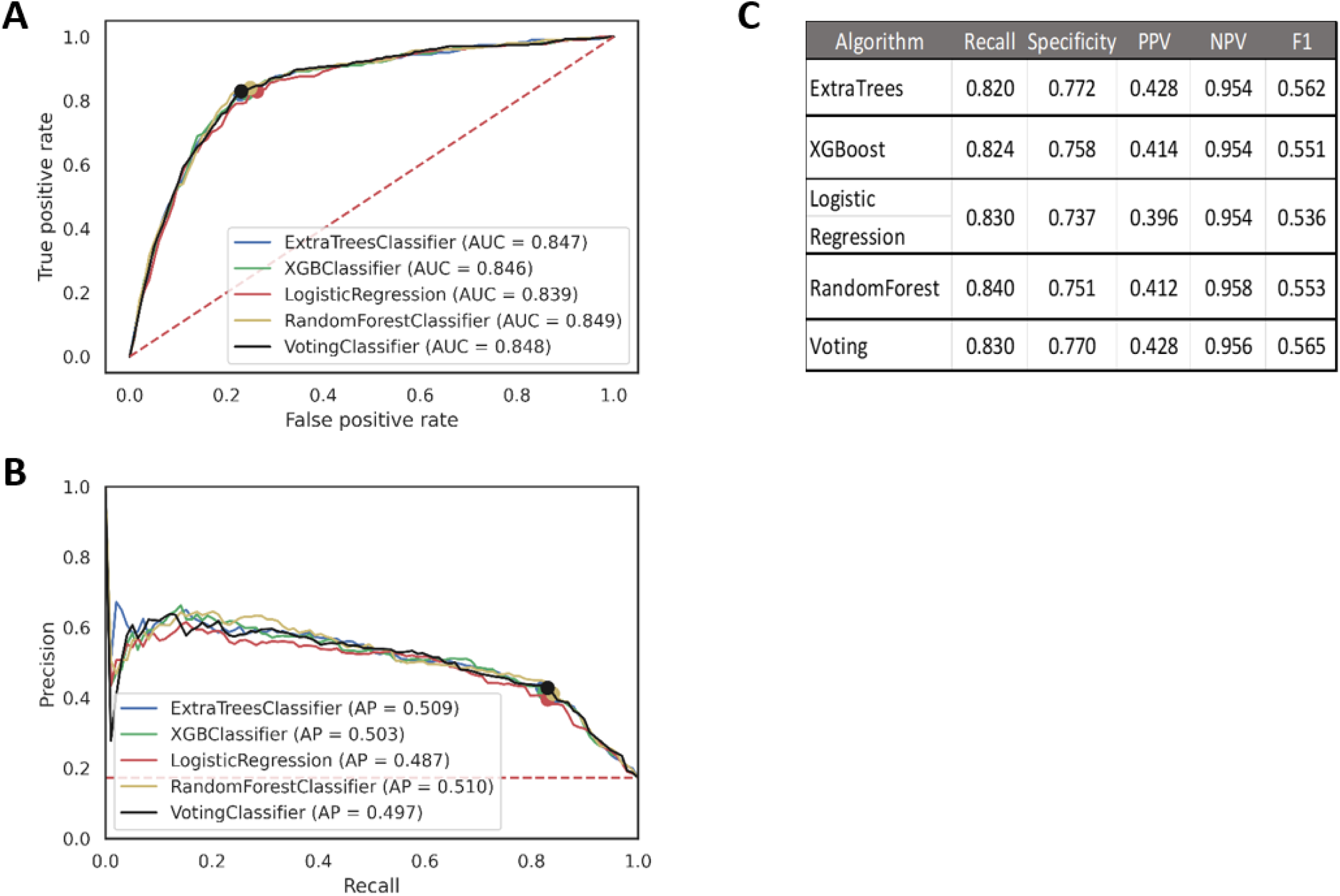
Full model performance. **A,** ROC curve on the test set for all classifiers. **B,** PR curve on the test set for all classifiers. **C,** Test set performance using operating threshold derived from validation sets during 5-fold cross validation.

Feature importance and model explainability were assessed through SHAP values from the training set predictions. Figure 3 displays the SHAP summary plot for the top 20 most important features in the full model. Features related with symptoms are regarded as the most important by the model, with asymptomatic, chills, cough, and headache are included in the top five. The model also successfully identifies that someone who is in self-isolation after COVID-19 exposure will have high risk of infection. Common COVID-19 symptoms such as fever, runny nose, sore throat, and muscle pain were also deemed important features by the model. Health workers that wore N95 masks, medical gloves, hazmat suits, and face shields regularly were also at lower risk of COVID-19 diagnosis. Furthermore, other behavioral features such as frequency and duration of outdoor activity, frequency of offline meetings, and density of people in the room most frequented by the health worker are also associated with higher COVID-19 risk. The high ranking of behavioral features highlights the benefit of adding these factors to complement symptomatic factors.

**Fig. 3.**
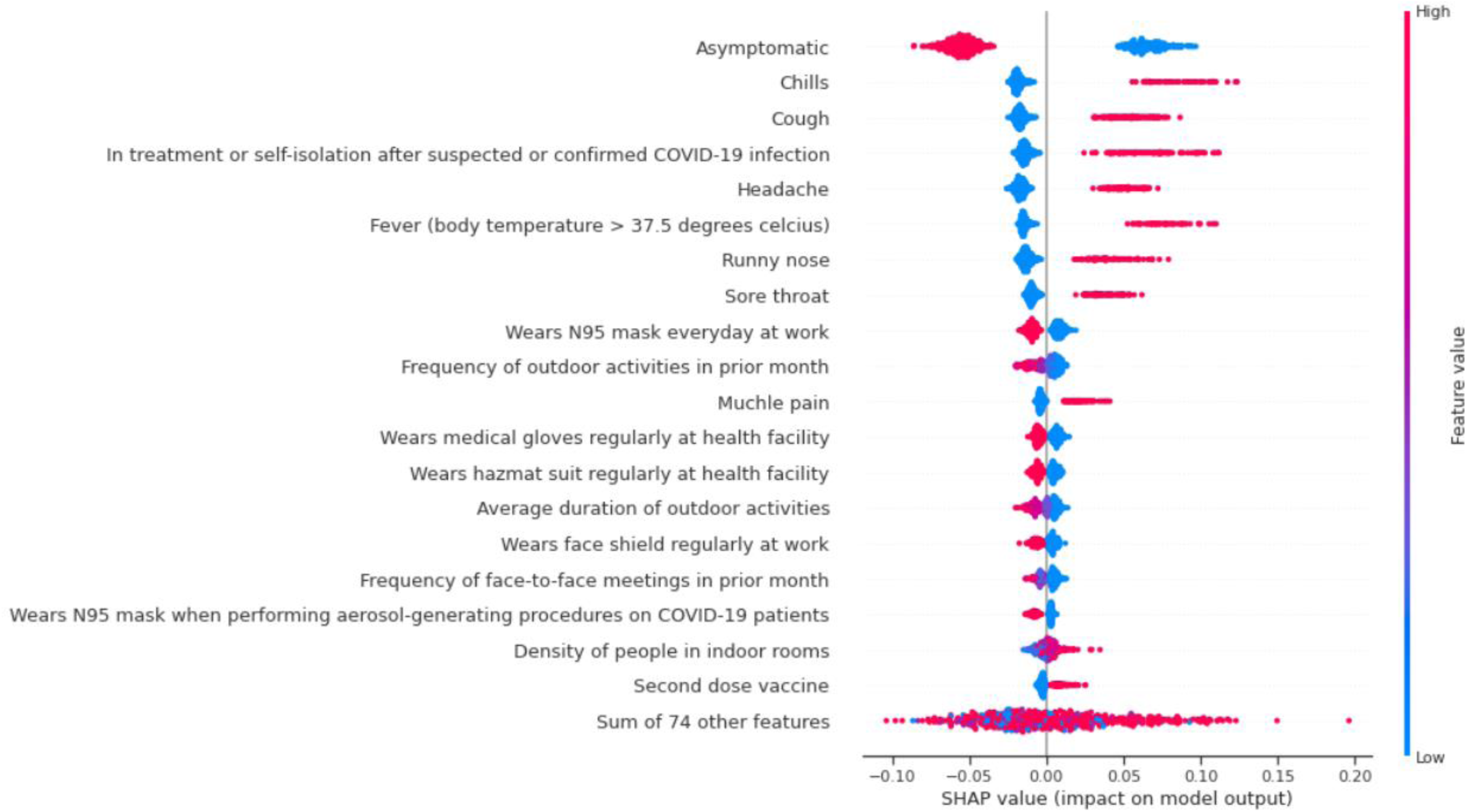
Full model explainability. SHAP analysis on training set predictions.

#### Jakarta Model

The Jakarta model was trained and cross-validated on health workers from Jakarta who submitted their results in 2021. Then, the model was tested on two datasets, which are data respondents from Semarang and Jakarta in 2022. The Semarang test set and Jakarta 2022 set was composed of 12.7% and 18.76% of the entire dataset respectively. The fraction of positive classes in the validation dataset was 10.27%, while the Semarang and Jakarta 2022 testing datasets were 42.2% and 31.3% respectively. As shown in Figure 4, the XGBoost classifier had the best predictive performance during cross-validation. The mean AUC (0.857 ± 0.017) outperformed random forest (0.856 ± 0.016), extra trees (0.856 ± 0.019), voting classifier (0,856 ± 0.017) and logistic regression (0.843 ± 0.015) (Fig. 4A). The random forest (0.434 ± 0.039) and extra trees (0.434 ± 0.052) produced the best AP, followed by voting classifier (0.429 ± 0.043), XGBoost (0.416 ± 0.045), and logistic regressor (0.409 ± 0.041) (Fig. 4B). The calibration curves from training showed that random forest is the most well-calibrated model, while the other models appear to have poorly calibrated predictions in upper predicted probability bins (Fig. 4C). Additionally, random forest had the lowest brier scores of 0.080 ± 0.0003.

**Fig. 4.**
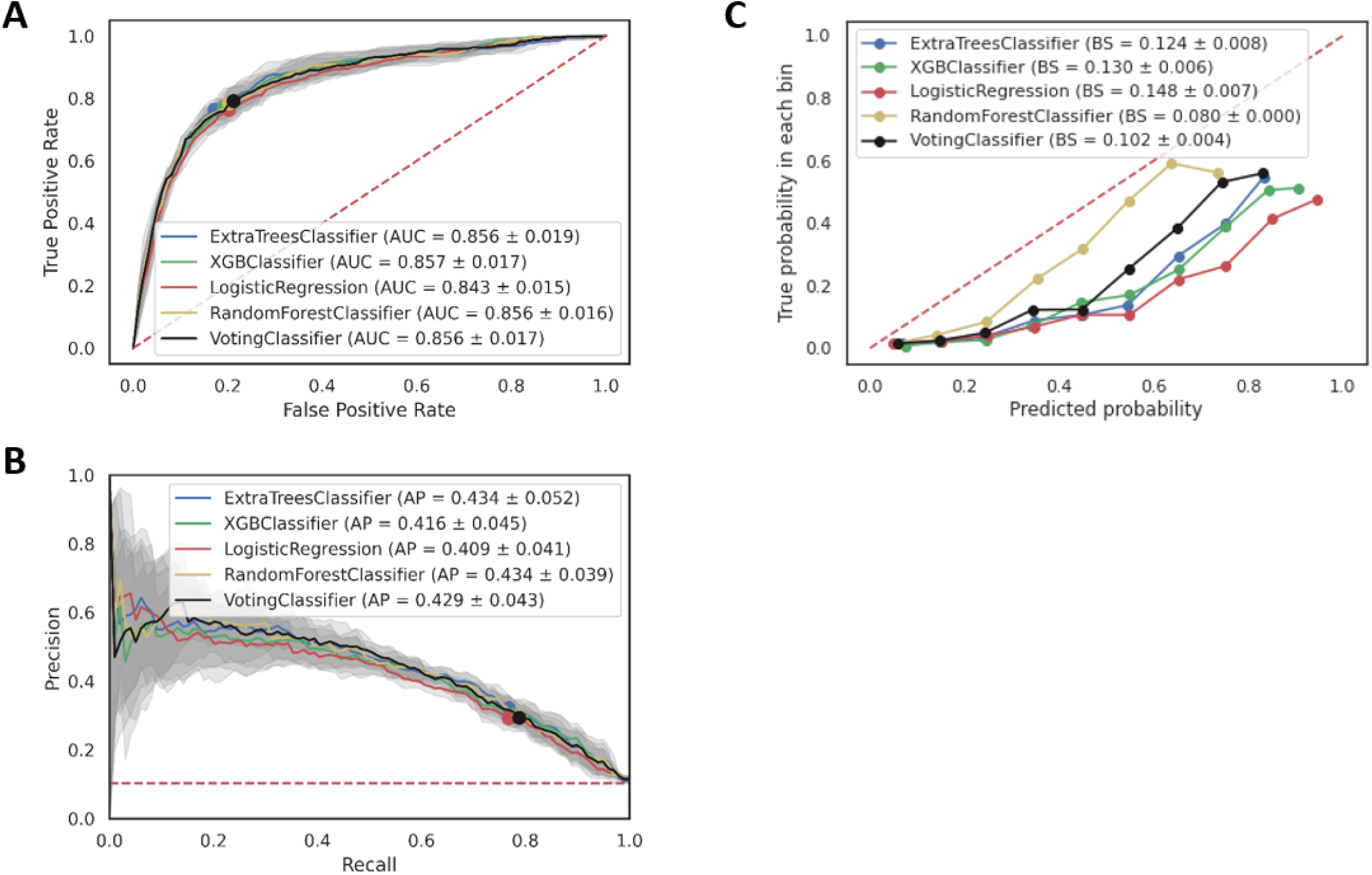
Jakarta model cross validation results. **A,** ROC curve on the validation sets for all classifiers. **B,** PR curve on validation sets for all classifiers. **C,** Calibration curve for all classifiers generated from validation sets during 5-folds cross validation.

When testing the models on the Semarang dataset, the random forest had the best AUC of 0.745 followed by extra trees, XGBoost, voting classifier, and logistic regression classifier with 0.744, 0.743, 0.740, and 0.726, respectively (Fig. 5A). XGBoost had the best AP of 0.705 followed by random forest (0.694), voting classifier (0.694), extra trees classifier (0.689) and logistic regression (0.672) (Fig. 5B). Looking at the F1 score, extra tree classifier got the highest score with 0.657 followed by logistic regression, random forest, XGBoost, and voting classifier with 0.651, 0.649, 0.646, and 0.646, respectively (Fig. 5C).

**Fig. 5.**
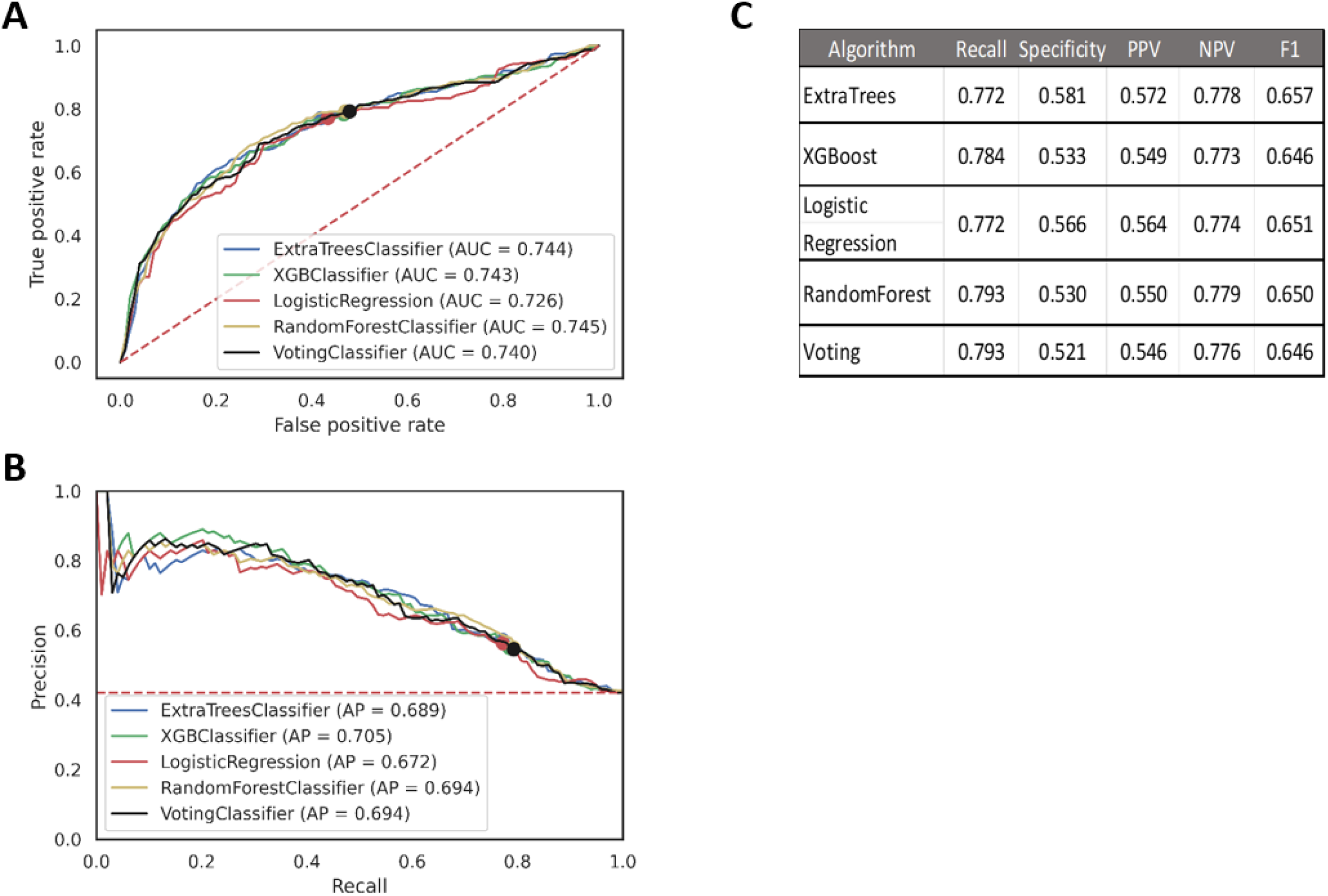
Jakarta model performance on Semarang dataset. **A,** ROC curve on the Semarang test set for all classifiers. **B,** PR curve on Semarang test set for all classifiers. **C,** Test set performance using operating threshold derived from validation sets during 5-fold cross validation.

On the other hand, testing on Jakarta 2022 dataset produced the best AUC of 0.762, which is associated with the voting classifier. Moreover, the random forest had a slight difference with AUC of 0.761. Meanwhile, XGBoost, extra trees, and logistic regression got 0.760, 0.757, and 0.753, respectively (Fig. 6A). The highest AP is achieved by voting classifier and logistic regression with 0.548 and 0.547. Furthermore, random forest, XGBoost, and extra trees followed with 0.535, 0.529, and 0.524 (Fig. 6B). The F1 score of random forest outperformed the other models with 0.582. The second and third places were occupied by XGBoost (0.502) and voting classifier (0.481) (Fig. 6C). Unfortunately, the F1 scores for this dataset experienced poorer performance compared to other test sets. It highlights timed-based drift in the data which influences the choice of threshold to be suboptimal [35].

**Fig 6.**
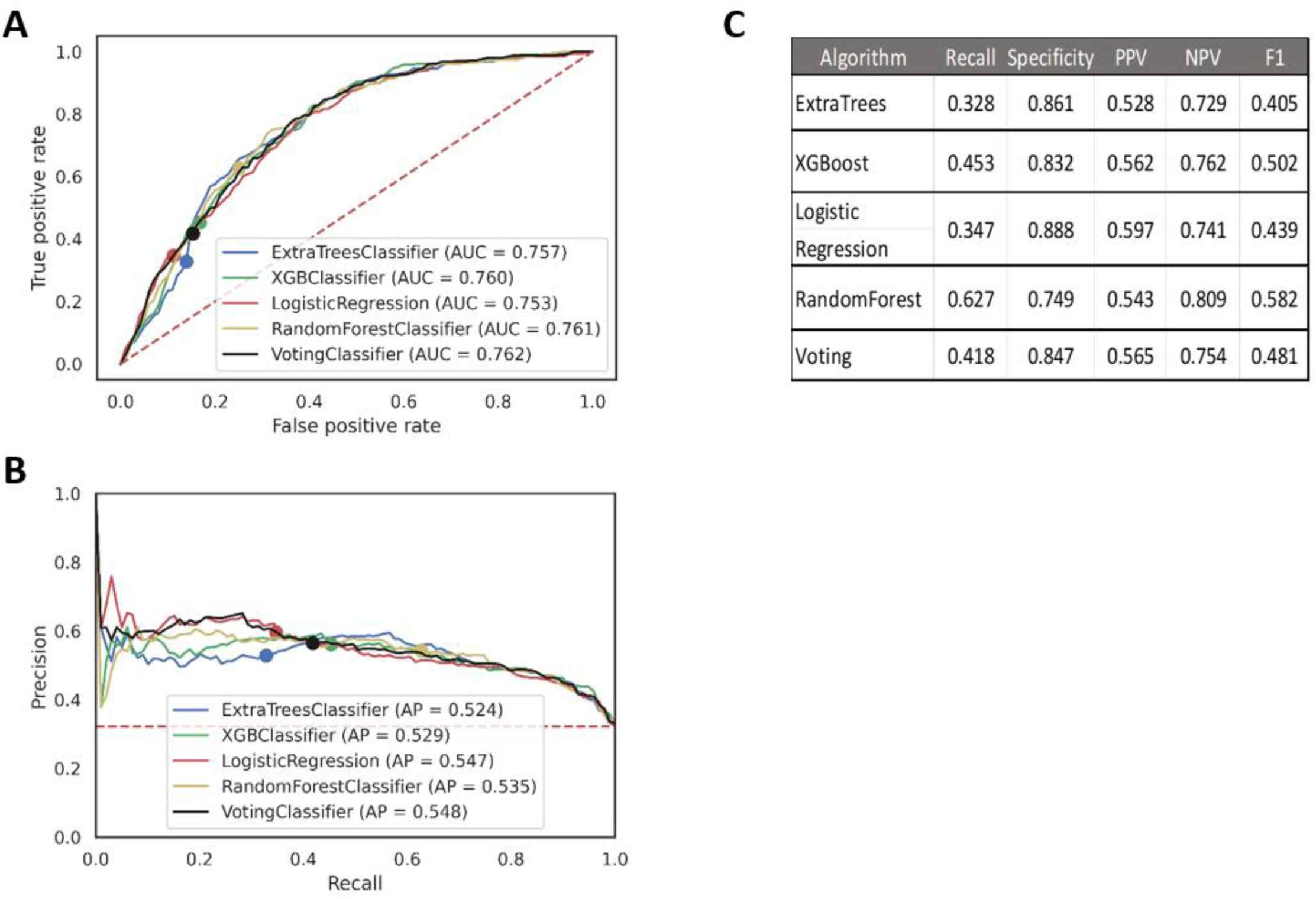
Jakarta model performance on Jakarta 2022 dataset. **A,** ROC curve on the Jakarta 2022 test set for all classifiers. **B,** PR curve on Jakarta 2022 test set for all classifiers. **C,** Test set performance using operating threshold derived from validation sets during 5-fold cross validation.

Based on the AUC, AP, and Brier Score, random forest classifier was chosen for the Jakarta model due to well-calibrated predictions and high training and testing performance. SHAP analysis is then executed using random forest as the model. Figure 7 displays a SHAP summary plot for the Jakarta model. The feature importance for the Jakarta model is highly identical with the full model. Almost half of the top 20 features are related to symptoms of COVID-19, which shows that symptoms are key to predicting infection. Moreover, outdoor activities also contribute to the risk of COVID-19 infection as shown by the high ranking of SHAP values of the average duration, frequency, and people density of outdoor activities. Similar to the full model, the model also picked wearing personal protective equipment (such as N95 mask, medical gloves, hazmat suit, and surgical hood) regularly as features which could influence the risk of infection. Additionally, the Jakarta model also recognized washing hands after wearing masks as a predictive feature for the model.

**Fig. 7.**
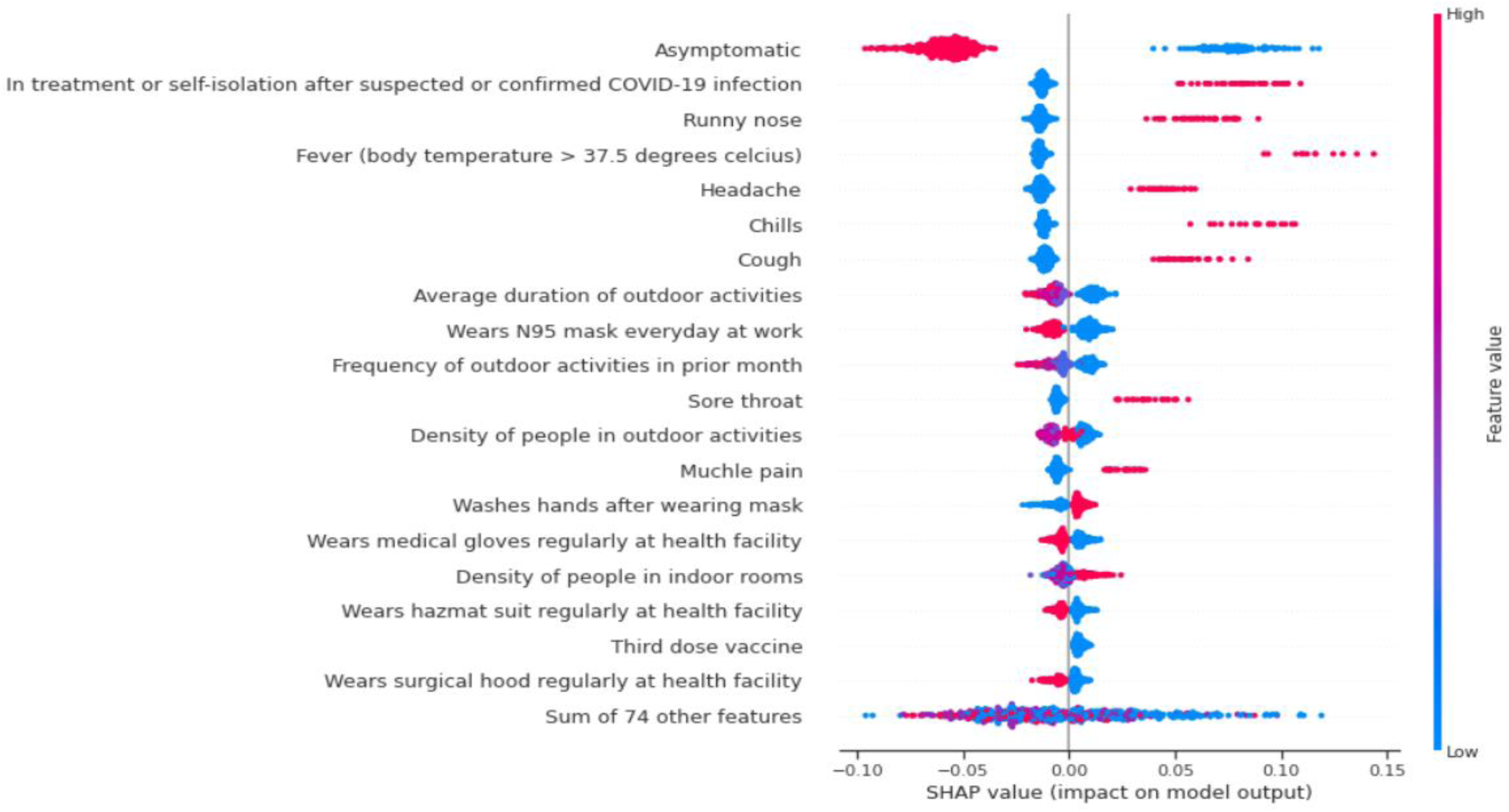
Jakarta model explainability. SHAP analysis on training set predictions.

## DISCUSSION

This study investigated the capability of information collected via questionnaires including sociodemographic information, behavioral tendencies, COVID-19 vaccine status, symptoms, comorbidities, and working conditions among others to predict COVID-19 diagnosis for Indonesian healthcare workers. We demonstrated that machine learning methods, specifically the random forest, XGBoost, logistic regression, extra trees, and ensemble algorithms of these models were able to predict COVID-19 diagnosis with a high performance. Models built on symptom data only have been found to be insufficient for application to clinical practice [37], perhaps due to the lack of sociodemographic, behavioral, comorbidity, or other critical risk factors for COVID-19 infection [38]. The models developed here incorporate many of these factors, and specifically include inputs on behavioral tendencies that are important in COVID-19 transmission and infection. The importance of behavioral tendencies is demonstrated by the SHAP summary plots for both models (Fig. 3 and Fig. 7), where behavioral tendencies ranked amongst the most important features for the models.

The full model performed well during training and testing with the random forest and was therefore chosen as the classifier of choice for the full model. Based on the AUC and AP, tree-based models (random forest, XGBoost, and extra trees) performed much better compared to linear models (logistic regression). One reason for this is because the tree-based models could extract non-linear relationships between the features and the target values, while logistic regression is restricted to linear patterns. Another advantage of tree-based models is their ability to capture the interaction between features and use those interactions as predictive signals. The outperformance of the tree-based model demonstrated the importance of non-linear relationship and feature interaction in predicting COVID-19 infection. Lastly, the tree-based models are an ensemble of decision trees which are aggregated to make a prediction. This method helps reduce the variance and avoid overfitting the training data.

A similar effect also can be observed in the Jakarta models, where the logistic regression had the worst AUC and AP than the other models in most cases. Random forest is also chosen as the final model for the Jakarta model due to its stability in performance and calibration. Additionally, the poorer performance of the Jakarta model on the testing Semarang healthcare worker dataset is likely due to potential drifts and biases in the testing data relative to the training data. This also happened in the Jakarta 2022 dataset, which suggests a time-based drift that changes the pattern in the data. One solution to this is to retrain the model periodically using the latest data. However, this approach will require a much larger dataset to build and evaluate a stable system.

Improved protective behavioral tendencies, such as wearing protective equipment regularly and limiting outdoor activities, as indicated by the models, are critical to the protection of healthcare workers during the pandemic [38]. The models we built are inclusive of many behavioral risk factors of COVID-19 infection among a myriad of other inputs and perform well on unseen data. Mass health worker testing, adequate PPE supply, self-isolation and quarantine, and education are the main recommendations previously issued for saving the frontline health workers during the pandemic [39]. CLM and our results may have the potential to assist in most of these guidelines, through assessing COVID-19 risk, prioritizing testing and thus isolation measures, and demonstrating the importance of utilizing PPE. Furthermore, CLM can be used to reduce costs, as the survey can be provided to health facilities at no cost.

Comparing the test and the training results for the Jakarta model, the test set AP is higher than the training set AP despite having a lower AUC. This is because the training dataset has a considerably smaller percentage of positive cases than the test datasets (Table 1). Since AP only focuses on the positive class, the baseline for AP is proportional to the percentage of positive cases. Therefore, a model used on a more balanced dataset will likely have a higher AP compared to an imbalanced one. On the other hand, both Semarang and Jakarta 2022 datasets have a more balanced distribution, causing a higher AP in the testing datasets.

This study has several limitations. The first limitation is the self-reported nature of the survey, which poses risks of over or underreporting. Methods of fraud detection and error handling must be applied if using the model in real-time. The authority for hospitals to recommend their staff to be included in the study may introduce selection bias in the data collection process. Additionally, recall bias may be introduced in answers to retrospective questions in the survey, such as symptoms within the previous 14 days. Future work for the study includes collecting more data for these models, as well as investigating models for using CLM survey and other data to predict additional outcomes, such as hospitalization and mortality. Recruitment of health workers for the study also will expand to several other provinces within Indonesia.

In April 2022, COVID-19 positivity and mortality rates started to decrease [40]. Consequently, the Indonesian government started to ease some of public health measures. With the loosening of public health measures, including COVID-19 test for travel clearance and use of mask in outdoor settings, our model can position itself in identifying healthcare workers at high risk of COVID-19 infection, allowing confirmatory testing and self-isolation. Further, it could promote early detection of COVID-19 infection and warn of any surge in cases to prevent outbreaks in healthcare facilities and anticipate a seasonal surge of COVID-19 cases [41]. Therefore, this model has the potential to facilitate surveillance in Indonesia as the Indonesian government has begun to transition into COVID-19 endemic phase [42] while the national vaccination rate for second dose of COVID-19 vaccine as of March 2023 is still at 74.51% [43].

## SUPPORTING INFORMATION

**S1 File. STROBE Checklist**

**S2 File. List of questions used in survey for healthcare workers**

## Data Availability

The datasets generated and analyzed in this study are not publicly available since it contains sensitive personal information, but are available from the corresponding author on reasonable request.

