## Supplemental File 2 for "Application of Machine Learning in Prediction of COVID-19 Diagnosis for Indonesian Healthcare Workers"

**S2 File. List of questions used in survey for healthcare workers**

**Question as Listed in Survey**

In the last 14 days, have you experienced any of the following symptoms?

How many days ago did you first started experiencing the symptoms?

Are you aware of the standard procedures involving personal protective equipment (PPE) according to the recommendations from the Ministry of Health?

In the last month, have you ever done any of the following without PPE for at least 15 minutes:

1. Touching,
2. Being within a distance of 1 meter,
3. Or processing specimens or cleaning contaminated equipment/room of a COVID-19 patient?

Are you currently in treatment or isolation after being in contact with a COVID-19 patient?

What is your role in the healthcare facility?

In the last month, what departments have you worked in?

In the last month, what PPE do you use most regularly in your daily work?

In the last month, have you ever conducted a procedure involving the release of aerosols from a COVID-19 patient?

examples given:

a) tracheal intubation;

b) non-invasive ventilation;

c) tracheostomy;

d) cardiopulmonary resuscitation;

e) manual ventilation before intubation;

f) taking swabs;

g) dental examination;

h) nose and throat examination;

i) other aerosol-involved procedures (such as nebulizer, bronchoschopy, etc.),

What PPE do you always use when conducting such procedures?

In the last month, do you regularly wear a mask when meeting someone not in your household?

In the last month, what type of mask do you use regularly in your daily activities?

In the last month, have you ever removed your mask partially or completely while meeting someone from outside your household?

In the last month, what position do you usually wear your mask?

In the last month, how many times have you been in a room with closed air circulation, other than your home (for examples, changing room, office, minimarket, or movie theater)?

How long did you stay in the room you visit most frequently?

What is the estimated average density of people in the room you visit most frequently?

In the last month, how many times have you attended an in-person social or professional gathering (for examples, family gathering, classroom, seminar, meetings, etc.)?

How long were those gatherings on average?

What was the estimated average density of people during those gatherings?

In the last month, how many times have you done an outdoor activity (for examples, jogging, biking, or working outside the office)?

What is the average duration of those activities?

What was the estimated average density of people during those activities?

In the last month, how many times did you use mass public transportation (for examples, train, or bus)?

How long were you in the vehicle on average?

What was the estimated average density in the vehicle?

In the last month, how many times did you use a ride-share, taxi, bajaj or similar modes of transportation?

How long were you in the vehicle on average?

In the last month, how many times did you fly on a commercial airplane?

How long were you in the airplane on average?

What was the estimated average density of people during your flight?

What type of mask do you wear during a flight?

Are you aware of the recommended six movements of handwashing?

In the last month, how often did you wash your hands involving those six movements?

(only ask if answered “Yes” from previous question)

When do you usually wash your hands using soap or hand sanitizer?

In the last month, did you avoid handshakes when meeting other people (other than family)?

Other than your responsibilities in your workplace, did you keep a minimum distance of one meter when meeting people from outside your household?

Do you still keep the minimum distance even when wearing a mask?

In the last month, has anyone from your household just returned from another country or city?

In the last month, have you visited another country or city?

How many days ago did you visit the country or city?

What mode of transportation did you use?

Please list the cities or countries you visited.

Are you currently diagnosed with a condition or infection?

In the last month how much did you weigh on average? (in kilograms)

What is your height? (in centimeters)

In the last month, did you smoke any tobacco products?

In the last month, how many cigarettes did you smoke weekly on average?

Did you smoke before last month?

Before your decision to quit smoking, how many cigarettes did you smoke weekly on average?

At what age did you quit smoking completely?

At what age did you quit smoking regularly?

Do you smoke e-cigarettes?

Age

Sex

Postal Code

What is the total income you and your partner earned in total last month?

How many dependents did you have in the last month? (including yourself)

What level of education did you achieve?

Are you vaccinated for COVID-19?

Which vaccine did you receive?

When did you receive your first dose?
